# Therapeutic application of alpha-1-antitrypsin in COVID-19

**DOI:** 10.1101/2021.04.02.21252580

**Authors:** Felix Ritzmann, Praneeth Chitirala, Yiwen Yao, Nadine Krüger, Markus Hoffmann, Wei Zuo, Frank Lammert, Sigrun Smola, Nastasja Seiwert, Naveh Tov, Noga Alagem, Bahareh Mozafari, Katharina Günther, Martina Seibert, Sabrina Hörsch, Thomas Volk, Philipp M. Lepper, Guy Danziger, Stefan Pöhlmann, Christoph Beisswenger, Christian Herr, Robert Bals

## Abstract

**Rationale:** The treatment options for COVID-19 patients are sparse and do not show sufficient efficacy. Alpha-1-antitrypsin (AAT) is a multi-functional host-defense protein with anti-proteolytic and anti-inflammatory activities.

**Objectives:** The aim of the present study was to evaluate whether AAT is a suitable candidate for treatment of COVID-19.

**Methods:** AAT and inflammatory markers were measured in the serum of COVID-19 patients. Human cell cultures were employed to determine the cell-based anti-protease activity of AAT and to test whether AAT inhibits the host cell entry of vesicular stomatitis virus (VSV) particles bearing the spike (S) protein of SARS-CoV-2 and the replication of authentic SARS-CoV-2. Inhaled and / or intravenous AAT was applied to nine patients with mild-to-moderate COVID-19.

**Measurements and Main Results:** The serum AAT concentration in COVID-19 patients was increased as compared to control patients. The relative AAT concentrations were decreased in severe COVID-19 or in non-survivors in ratio to inflammatory blood biomarkers. AAT inhibited serine protease activity in human cell cultures, the uptake of VSV-S into airway cell lines and the replication of SARS-CoV-2 in human lung organoids. All patients, who received AAT, survived and showed decreasing respiratory distress, inflammatory markers, and viral load.

**Conclusion:** AAT has anti-SARS-CoV-2 activity in human cell models, is well tolerated in patients with COVID-19 and together with its anti-inflammatory properties might be a good candidate for treatment of COVID-19.

**Funding:** This work was supported by grants of the Rolf M. Schwiete Stiftung, the Saarland University, the BMBF, the State of Lower Saxony, and The State of Saarland.

**Scientific Knowledge on the Subject:** COVID-19 is caused by “severe acute respiratory syndrome coronavirus 2” (SARS-CoV-2) and is a serious global health threat. Efficacious treatments are not available and there are no drugs that can prevent progression towards respiratory and extra-pulmonary organ failure. AAT has been studied *in vitro* and has activity against SARS-CoV-2. We searched PubMed and Google Scholar using the search terms “COVID-19”, “SARS-CoV-2”, “therapy”, and “α-1-antitrypsin” (AAT) for research published in 2020 and 2021.

**What This Study Adds to the Field:** This study shows the results of a translational program with a focus on the biology of AAT in COVID-19. The data show that there is a relative deficiency of AAT in relation to systemic inflammation. AAT inhibits serine protease activity in human airway cells and the replication of SARS-CoV-2 in human lung organoids. Inhaled and / or intravenous application of AAT in nine patients was associated with clinical stabilization. The findings of this exploratory study suggest that AAT has a mechanistic role in the pathophysiology of COVID-19 based on its anti-inflammatory and anti-viral activities. This offers the possibility to test and develop AAT application for treatment of different phenotypes or stages of COVID-19, including severe, inflammatory courses or early stages. Inhaled treatment could be an option to administer AAT non-invasively in early stages.

## Introduction

Coronavirus disease 2019 (COVID-19) is a novel illness and is a rapidly evolving pandemic with unprecedented impact on global health in the last 100 years. COVID-19 is caused by “severe acute respiratory syndrome coronavirus 2” (SARS-CoV-2), a member of the *Coronaviridae* family that is regarded a spill-over of an animal coronavirus to humans (1). At the time of submission of this work, SARS-CoV-2 has caused 80 million diagnosed infections and 1·8 million known deaths. The trajectories of the pandemic are still difficult to predict. While vaccines are currently developed and enter public health prevention measures, the treatment options for COVID-19 patients are sparse and do not show sufficient efficacy. Remdesivir (2) and dexamethasone (3) are used in hospitalized patients and use of tocilizumab has also shown beneficial outcomes for this patient group (4). The options for patients with mild COVID-19 or asymptomatic infection are scant. At the present time, there are no efficacious drugs available that can be applied non-invasively and decrease the likelihood of disease progression towards organ failure (5).

SARS-CoV-2 uses several host factors to infect target cell including the transmembrane protease serine 2 (TMPRSS2) for activation of the viral spike (S) protein, and angiotensin-converting enzyme 2 (ACE2) for entry (6-8). The disease pathomechanisms comprise the direct consequences of cell and tissue damage and a systemic inflammatory syndrome that includes innate and adaptive immune activation, hypercoagulation and other features (9, 10).

Alpha-1-antitrypsin (AAT) is a multi-functional host-defense and acute phase protein and member of the serpin superfamily (11). AAT is mainly produced in the liver and secreted into the blood stream. In addition, AAT is synthesized in macrophages and lung epithelial cells. Genetic deficiency results in increased susceptibility for early chronic obstructive lung disease (COPD) or chronic liver injury (11). AAT has a broad anti-protease activity and has been implicated in several biological processes such as the regulation of inflammation (12, 13). Intravenous or inhaled AAT has been used in multiple clinical trials for COPD or cystic fibrosis and has a good safety profile (14, 15). In the context of hepatitis C virus infection, it has been shown that AAT inhibits TMPRSS2 (16), the same host serine protease that is necessary for the activation of the S-protein of SARS-CoV-2 (6).

Based on potential inhibition of virus entry by AAT and its broad anti-inflammatory activity, the aim of this study was to investigate whether AAT can be considered as candidate for the treatment of COVID-19.

## Materials and methods

### Methods

#### Patients for biomarker measurements

Patients for observational measurements of AAT serum levels were recruited within the COVID-19 cohort study CORSAAR (n = 35) and its control cohort PULMOHOM. Both studies have been approved by the ethics committee of the Landesärztekammer Saarland, and all patients or their legal representatives gave their informed consent. Serum C-reactive protein (CRP), AAT, and interleukin-6 (IL-6) levels were measured by multiplex arrays. Moderate disease severity was defined as hospitalized patients without need for intensive care treatment, severe disease was defined as patients with need for intensive care treatment.

#### Reagents

AAT (A6150), aprotinin (trasylol) (A3428), and camostat mesylate (SML0057) were purchased from Merck (Germany). AAT was additionally obtained from Kamada (Rehovot, Israel). Camostat mesylate (Tocris, cat. no. 3193) was reconstituted in dimethyl sulfoxide (DMSO), yielding a stock concentration of 10 mM.

#### Cell culture

HEK-293T (human embryonic kidney; DSMZ no. ACC 635) cells were cultivated in Dulbecco’s modified Eagle medium containing 10 % fetal bovine serum (FCS, Biochrom), 100 U/ml of penicillin and 0.1 mg/ml of streptomycin (PAN-Biotech). Calu-3 cells (human lung adenocarcinoma; kindly provided by Stephan Ludwig, Institute of Virology, University of Münster, Germany) were cultivated in minimum essential medium supplemented with 10 % FCS, 100 U/ml of penicillin and 0.1 mg/ml of streptomycin (PAN-Biotech), 1x non-essential amino acid solution (from 100x stock, PAA) and 1 mM sodium pyruvate (Thermo Fisher Scientific). 16-HBE cells, a SV40-immortalized human bronchial epithelial cell line, were grown in Opti-MEM with 10 % FCS, penicillin, and streptomycin. All cell lines were incubated at 37 °C in a humidified atmosphere with 5 % CO2. HEK-293T were transfected by calcium-phosphate precipitation.

Airway basal cells were isolated and cultured as described before (17). Briefly, basal cells from the donors were cultured using Airway Epithelial Cell Growth Medium (C-21060; PromoCell) supplemented with 1 µM of A83-01 (TGFβ inhibitor) (2939; Biotechne), 0.2 µM of DMH-1 (BMP4 inhibitor) (4126; Biotechne) and 5 µM of Y27632 (ROCK inhibitor) (1254; Biotechne) to preserve prolonged basal cell phenotype with high proliferative capacity. Human lung organoids were cultured as described before (17). Human bronchial epithelial cells (HBECs) were isolated from fresh brush biopsies from patients who underwent bronchoscopy. Cells were washed from the brushes with AEC media (PromoCell, Germany) and expanded in conventional 2D co-culture on NIH-3T3 feeder cells and presence of 10 µM RHO/ROCK Inhibitor (Y-27632, StemCell) to avoid loss of differentiation capacity. For organoid culture, cells were harvested and transferred in differentiation media (BEGM, Lonza, with added supplements except for retinoic acid and T3, mixed 1:1 with DMEM-F12). A 96-well plate was precoated with 40 µl differentiation media containing 25% Matrigel. 80 µl of a cell suspension with 3 × 10^4^ cells/ml in differentiation media and 5% Matrigel was plated in each well. 120 µl differentiation media with 5% Matrigel was added on top on day 2 and changed every 4 days. The organoids were used for the experiments after 21 days of maturation. For this purpose, 120 µl of medium was removed and the Matrigel was dissolved by incubation with Cell Recovery Solution (Corning, USA). The organoids were then collected and washed with PBS. 100 organoids in 100 µl OptiMEM (Gibco) were transferred in each well of a black 96-well plate.

For immunohistochemistry, maturated organoids were harvested, washed in PBS, fixed in buffered 4% formaldehyde solution for 1 hour and embedded in paraffin. Deparaffinized paraffin sections (2 µm thickness) were treated with PBS with 1% BSA and 0.1% Tween-20 and incubated overnight with primary antibodies against TMPRSS2 and ACEII (Abcam #ab92323; R&D #AF933). Sections were incubated with appropriate HRP-conjugated secondary antibodies for 30 min at RT (Histofine, Nichirei Biosciences). Color reaction was developed using aminoethyl carbazole (AEC) chromogen.

The use of human cells was approved by the ethics committee of the Landesärztekammer of the Saarland and informed consent was obtained from the patients.

### Assessment of AAT anti-protease activity

The effect of AAT on epithelial cell trypsin-like protease activity was assessed using the synthetic peptide analogue (trypsin substrate), Boc-Gln-Ala-Arg-AMC (3135-V; PeptaNova GmbH, Germany). 16-HBE cell or organoids in 96 well plates were incubated with trasylol (100 or 200 µg / ml), AAT (1or 2 mg / ml), or camostat (250 µM). Finally, 2 µl of 100 µM peptide substrate was added to cells and fluorescence was measured immediately at 360 / 465 mm of excitation/emission wavelength every 2 minutes for 6 hours at 37°C. Trypsin-EDTA was used as positive control. All the compounds were dissolved in Opti-MEM.

### Viral infection models

Expression plasmids for severe acute respiratory syndrome coronavirus 2 spike glycoprotein (SARS-2-S) and vesicular stomatitis virus glycoprotein (VSV-G) have been described elsewhere (6). In addition, we used empty pCG1 expression plasmid that was kindly provided by Roberto Cattaneo (Mayo Clinic College of Medicine, Rochester, MN, USA). VSV pseudotype particles bearing SARS-2-S, VSV-G or no viral protein (negative control) were produced using an established protocol (18): First, HEK-293T cells were transfected to express the respective viral protein or no viral protein. At 24 h posttransfection, cells were inoculated with a replication-deficient VSV vector, VSV*ΔG-FLuc, that was kindly provided by Gert Zimmer (Institute of Virology and Immunology, Mittelhäusern, Switzerland)(19). The VSV vector lacks the genetic information for VSV-G and instead contains open reading frames for two reporter proteins, enhanced green fluorescent protein and firefly luciferase (FLuc). At 1 h postinoculation, cells were washed and fresh culture medium was added. For SARS-2-S expressing cells as well as cells expressing no viral glycoprotein, anti-VSV-G antibody (culture supernatant from I1-hybridoma cells; ATCC no. CRL-2700) was added to the culture medium. At 16-18 h postinoculation, VSV pseudotype particles were harvested. For this, the culture supernatant was collected and centrifuged (2,000 x g, 10 min, room temperature) to remove cell debris. Finally, the clarified supernatants were aliquoted and stored at −80 °C until further use.

For transduction experiments, Calu-3 cells (grown in 96-well plates) were preincubated with different concentrations of AAT (Kamada, diluted in culture medium) or culture medium alone. Following an incubation phase of 1 h at 37 °C, VSV pseudotypes bearing either VSV-G, SARS-2-S or no viral glycoprotein (control) were added to the cells, which were further incubated for 16-18 h, before transduction efficiency was analyzed. For this, the culture supernatant was removed and lysis buffer (1x Cell Culture Lysis Reagent, produced from 5-fold concentrated stock; Promega) was added. After 30 min of incubation at room temperature, cell lysates were transferred into white 96-well plates and samples were analyzed for FLuc activity using a commercial substrate (Beetle-Juice, PJK) and a Hidex Sense plate luminometer (Hidex).

Human organoids were pre-incubated with AAT (Kamada, final concentration: 5 or 10 mg/ml) or camostat mesylate (final concentration: 50 µM) or Opti-MEM containing no inhibitors for 2 h at 37 °C and 5 % CO2. Thereafter, organoids were infected with 1 × 10^5^ infectious units of SARS-CoV-2 isolate NK (isolated from an infected traveler retourning from Ischgl, Austria; kindly provided by Stephan Ludwig, Institute of Virology, University of Münster, Germany) for 1 h at 37 °C and 5 % CO2. The virus-containing inoculum was removed and organoids were washed carefully with PBS three times before 200 µl of Opti-MEM containing the inhibitors was added. At 1 h (washing control) and 24 h (end point) post infection (p.i.), 100 µl supernatant were collected and stored at −80 °C for virus titration. For virus titration, Vero E6 cells were grown in 12-well plates until reaching confluency. Cells were inoculated with 850 µl of serial dilutions of the collected supernatants and incubated for 1 h at 37 °C and 5 % CO2. The virus inoculum was removed, cells were washed with PBS and overlaid with 1 % plaque agarose (Biozym) dissolved in Eagle’s minimal essential medium without phenol red (Lonza). Plaques were counted at 3 d p.i. and viral titers were determined as plaque forming units (PFU)/ml.

### AAT treatment of patients with COVID-19

Based on its anti-viral and anti-inflammatory activities together with large experience from therapeutic applications of AAT and the low number of side effects of treatment, we decided to offer patients with moderate COVID-19 inhaled or intravenous AAT as individual medication. As a consequence, the focus of this approach was the care for the individual patient and not a controlled clinical trial. This approach was reported to the ethics committee of the Ärztekammer des Saarlandes. As inhaled drug we used nebulized Prolastin, which is a medicinal product nationally licensed for treatment of AAT-deficiency dependent lung disease. Prolastin has been used for inhaled treatment in several trials earlier (20). In addition, we used AAT (Kamada) for intravenous and inhaled use, which has been applied in clinical trials. This drug was imported after approval by the Ministerium für Soziales, Gesundheit, Frauen und Familie (Ministry of Social Affairs, Health, Women and Family) of Saarland. Four patients were treated with daily inhalation of 100 mg Prolastin over 15 min for a maximum of 7 days. Five patients were treated with combined intravenous and inhaled treatment with daily inhalation of 100 mg and infusion of 60 mg/kg body weight at day 1, 3 and 5. All patients received standard treatment for COVID-19 and associated complications. Viral load was measured in respiratory specimens using the cobas® SARS-CoV-2 Test, Roche Diagnostics, Switzerland or the RealStar® SARS-CoV-2 RT-PCR Kit 1.0 (altona diagnostics, Hamburg, Germany).

### Statistics

Continuous data are presented as mean +/-SD (standard deviation) when normally distributed, if not otherwise indicated. Categorical data are presented as percentages. Statistical differences between continuous variables were determined using Student’s *t*-test or chi-squared test for categorical variables. Differences between more than two groups were analyzed with ANOVA with Tukey post-hoc testing. Data were analyzed with Microsoft Excel (Microsoft, Redmond, USA), SPSS 28 (IBM, Ehningen, Germany) and graphed using GraphPad Prism 8 Software (San Diego, USA). The P level for significance was 0.05.

## Results

### Relative deficiency of AAT in severe COVID-19

AAT is an acute phase protein that has anti-protease and anti-inflammatory properties. Earlier results from COVID-19 patients indicated a relative deficiency of AAT in relation to inflammatory markers (21). We determined the AAT serum concentration and found that patients with COVID-19 have increased levels in the blood as compared to patients undergoing elective surgery for extra-pulmonary disease (Fig. 1A). Within the COVID-19 group, there were no differences of the AAT serum concentration between severe cases vs. moderate cases or survivors vs. non-survivors (data not shown). In the present study, the ratio IL-6 / AAT was significantly increased in non-survivors (Fig. 1B), The ratio CRP / AAT was significantly increased in patients with more severe disease (Fig. 1C). These data indicate that in COVID-19 there is a relative AAT deficiency with inadequate production of AAT in response to systemic inflammation.

**Figure 1:**
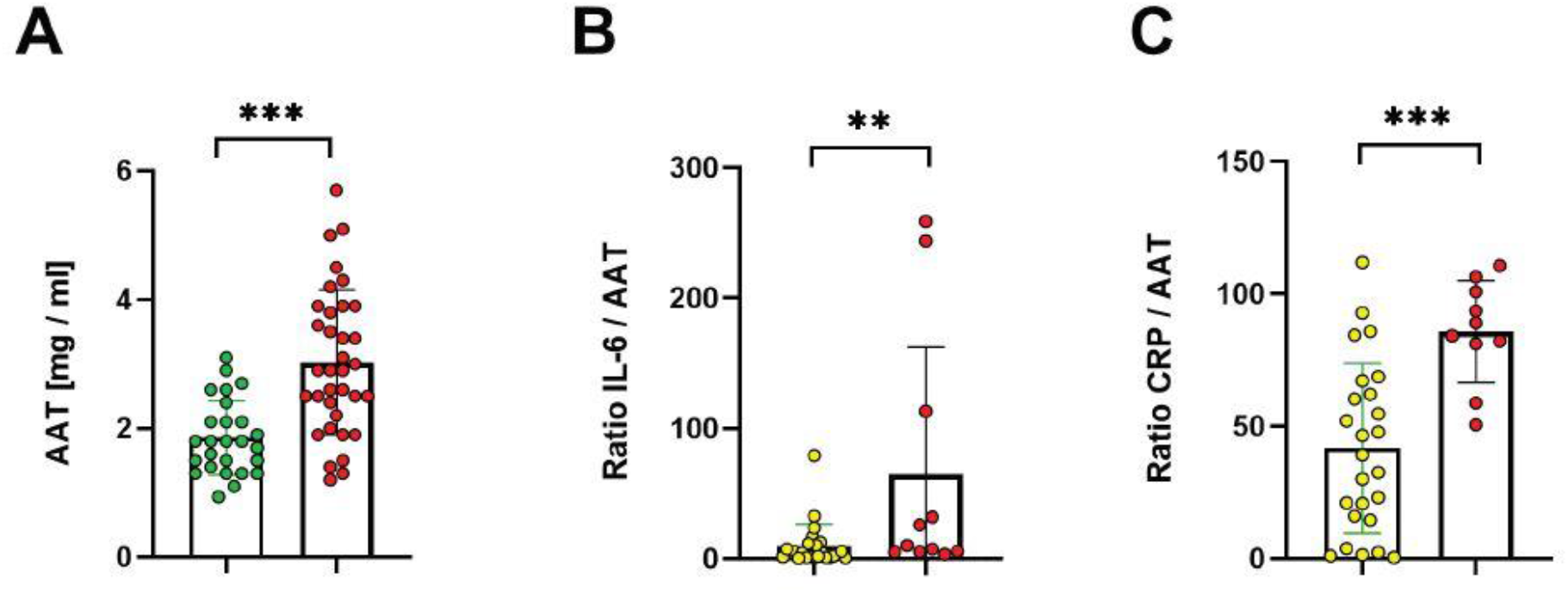
Relative deficiency of serum AAT in severe COVID-19. (A) Serum AAT concentration are increased in COVID-19 (red dots) patients as compared to controls (green dots) undergoing elective surgery (*t*-test). The ratios IL-6 / AAT and CRP / AAT are significantly increased in severe COVID-19 (B, red dots) or non-survivors (C, red dots), respectively (yellow dots in B: non-severe COVID-19, in C: survivors). Student’s t-test, data are shown as the mean ± SD with ** = p < 0.01 and *** = p < 0.001.

### ACE2 and TMPRSS2 are expressed in epithelial cells and human airway organoids

3D organoid cultures of airway epithelium were developed for high-throughput screening to identify possible TMPRSS2 inhibitors. To investigate whether TMPRSS2 and ACE2 (6) are expressed in epithelial cells of the organoids, immunostaining was performed and showed expression of both TMPRSS2 and ACE2 (Fig. 2A).

**Figure 2:**
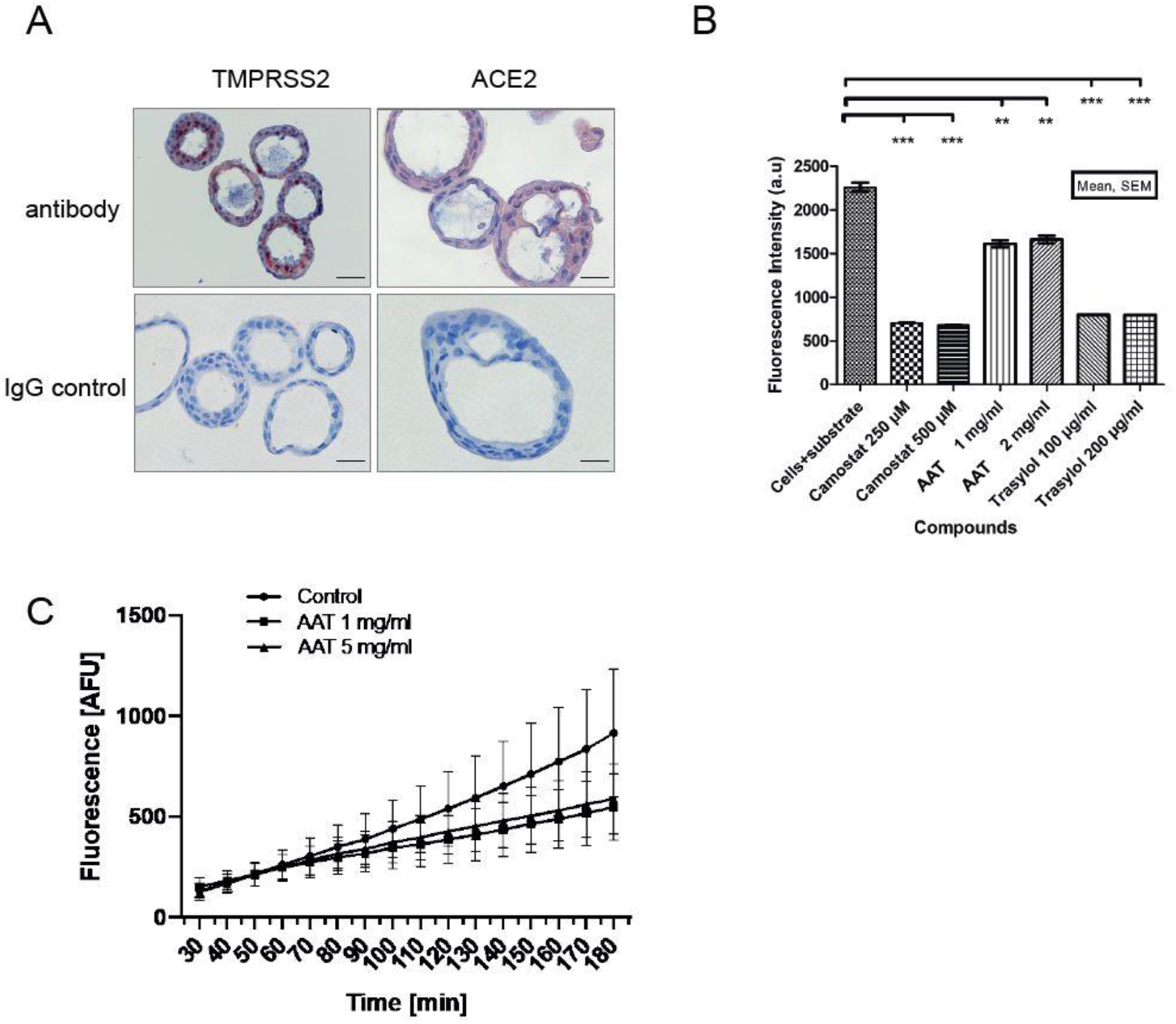
Human airway organoids as models to study SARS-Cov-2 infection. (A) TMPRSS2 and ACE2 are expressed in airway organoids as detected by immunohistochemistry. Representative images of organoids stained for TMPRSS2 and ACE2 with corresponding IgG controls. Scale bar 40 µm in TMPRSS2 column and 20 µm in the ACE2 column. (B) AAT and chemical compounds inhibit protease activity in airway epithelial cells using the peptide Boc-Gln-Ala-Arg-AMC as substrate. (C) AAT blocks the proteolysis of the synthetic peptide Boc-Gln-Ala-Arg-AMC in airway organoids. The slope of non-treated organoids and AAT treated organoids shows the time course of peptide proteolysis. Data from three independent experiments. ANOVA with Tukey post-hoc test. Data are shown as the mean ± SD with **p < 0.01 and ***p < 0.001.

### AAT inhibits protease activity in epithelial cells

Next, we analyzed the effect of protease inhibitors on the activity of trypsin-like proteases, including TMPRSS2, in in 2D and 3D cultures of airway epithelial cells using a fluorescence-based assay. The protease activity was measured by incubating cell cultures with the synthetic peptide analogue Boc-Gln-Ala-Arg-7-Amino-4-methylcoumarin, a substrate for trypsin-like proteases, including TMPRSS2. We found that AAT inhibits the protease activity in a concentration between 1 and 5 mg/ml in submersed, undifferentiated epithelial cells (Fig. 2B) or organoids (Fig. 2C). Camostat and trasylol were more active in cell lines (Fig. 2B).

### AAT inhibits the replication of SARS-CoV-2 in human airway organoids

Next, we investigated whether AAT inhibits SARS-CoV-2 spike protein (SARS-2-S)-driven entry into the human lung cell line Calu-3, which expresses endogenous ACE2 and TMPRSS2. For this, we employed previously described vesicular stomatitis virus (VSV)-based vectors bearing either SARS-2-S or the glycoprotein of VSV (VSV-G) (6). The protease inhibitor camostat, which is known to block TMPRSS2, was included as positive control in these experiments. Camostat reduced SARS-2-S but not VSV-G-driven entry efficiently and in a dose-dependent manner, as expected (6). Inhibition by AAT was also observed but was modest and only detected at high concentration (Fig. 3A). At 20 mg/ml a slight, unspecific inhibition of VSV-G-driven entry was detected. We next analyzed whether these results translate to human lung organoids and authentic SARS-CoV-2. Camostat efficiently inhibited SARS-CoV-2 infection of lung organoids, while an approximately 50% reduction of SARS-CoV-2 infection was observed at 10 mg/ml of AAT (Fig. 3B). These results suggest that AAT exerts moderate antiviral effects in human lung cells.

**Figure 3:**
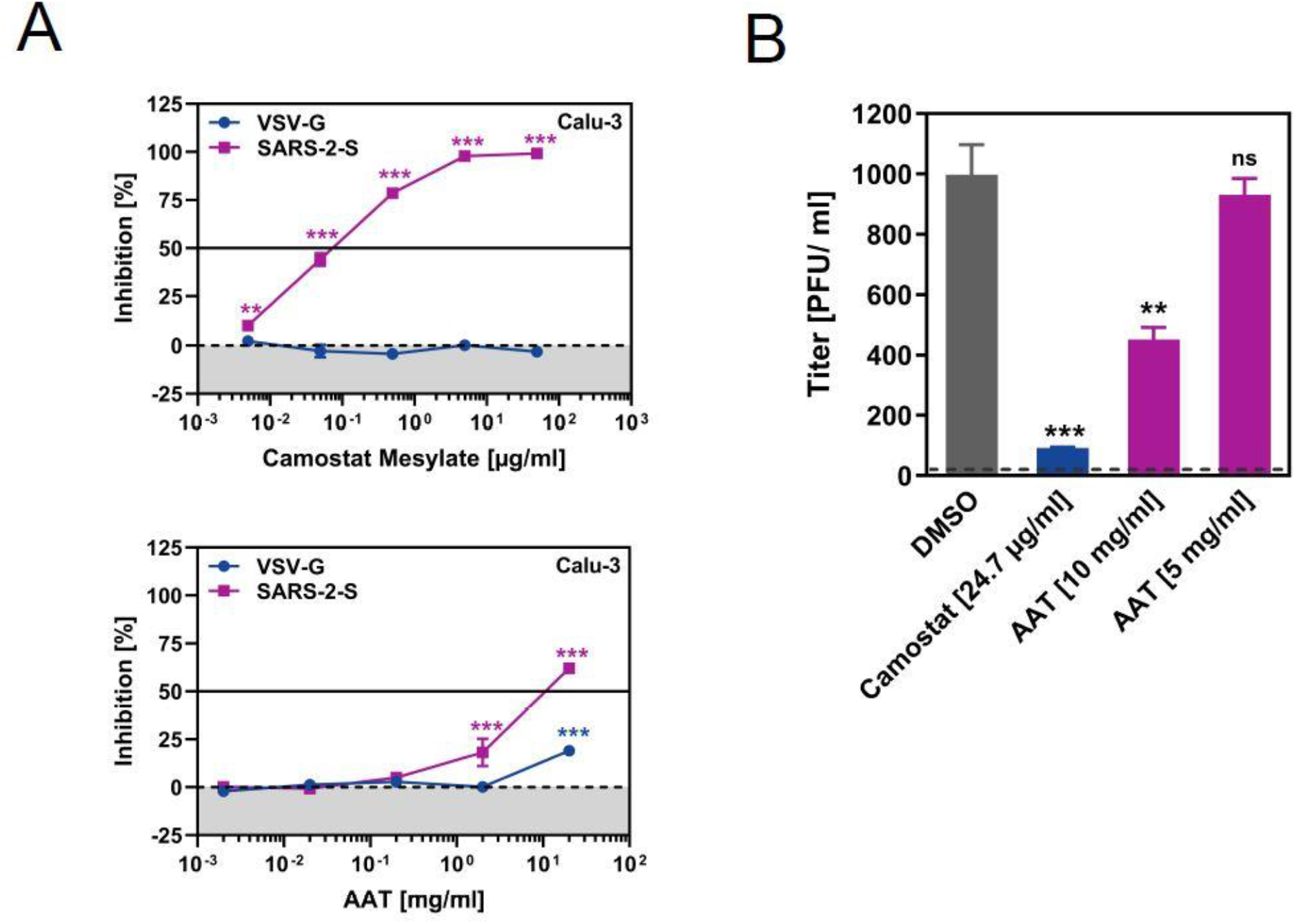
AAT exerts moderate antiviral activity. A) Calu-3 cells were preincubated for 1 h with the indicated concentrations of camostat or AAT and then transduced with VSV particles bearing SARS-2-S or VSV-G. Transduction efficiency was determined at 16 h post inoculation by measuring luciferase activities in cell lysates. The average of three independent experiments performed with technical quadruplicates is shown. Error bars indicate SEM. B) Human lung organoids were pretreated for 2 h with the indicated concentrations of camostat or AAT, inoculated with SARS-CoV-2 and viral titers determined at 24 h post infection using plaque assay. The average of two independent experiments performed with technical quintuplicates is shown. Error bars indicate SD with **p < 0.01 and ***p < 0.001.

### Application of AAT in patients

Based on the anti-viral and anti-inflammatory activities of AAT, we offered inhaled or combined inhaled / intravenous application to patients with mild to moderate COVID-19. Four patients were treated with inhaled AAT (100 mg/day for 7 days) and five patients were treated with inhaled (100 mg/day for 7 days) and intravenous treatment (60 mg/kg body weight, day 1,3,5). For further analysis we matched two COVID-19 patients from the CORSAAR cohort based on age and disease severity (patient characteristic in Table S1).

All patients treated with AAT survived and were discharged from the hospital in good functional status. To monitor disease activity, we recorded clinical measurements, inflammatory blood parameters and viral load. The respiratory status eventually improved in all patients; three patients experienced an initial deterioration (Fig. 4A). Serum CRP levels decreased over the days after initiation of the AAT application with individual variations (Fig. 4B). Virus load was monitored at irregular intervals and turned negative in all patients (Fig. 4C). We did not observe any drug-associated side effects such as allergic reactions. In the group of matched patients, three patients died, the decrease of CRP was delayed as compared to the AAT treatment group (Fig. 4B).

**Figure 4:**
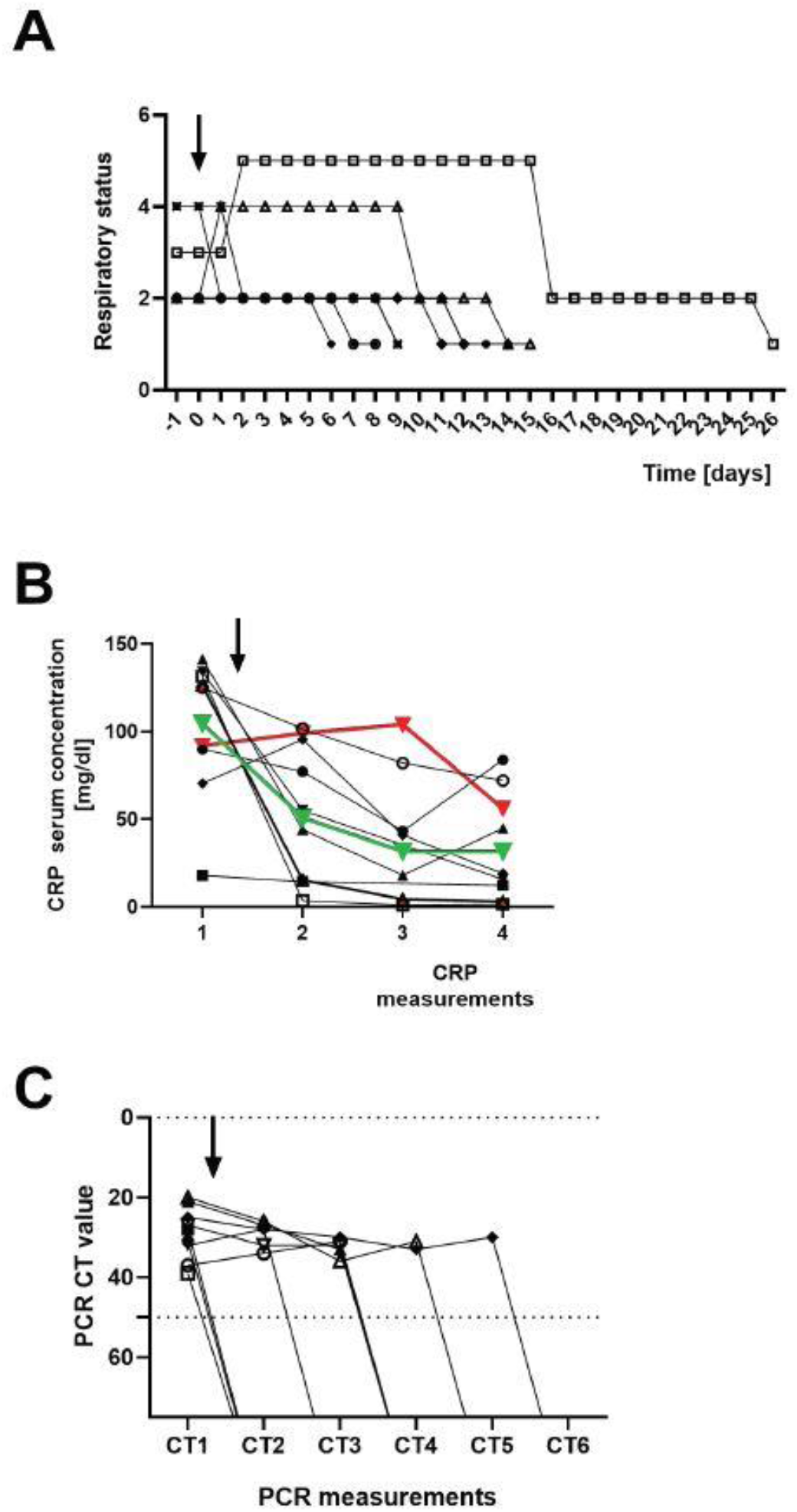
Application of AAT to patients with COVID-19. A) Development of the respiratory status in all 9 patients after the initiation of AAT treatment over time (arrow, respiratory status: 1 = spontaneous, 2 = oxygen, 3 = NIV, 4 = high flow oxygen, 5 = invasive, 6 = ECMO). B) Overall CRP serum concentrations were decreasing after AAT application (arrow). The green line represents the mean of all 9 patients with AAT reatment, the red line represents the mean of 18 matched control COVID-19 patients. C) The viral load in oro- or nasopharyngeal swabs decreased over time.

## Discussion

The main finding of this work is that AAT is a candidate for treatment of COVID-19. Evidence from this work and from the literature shows that AAT has anti-viral and anti-inflammatory activity. Administration of AAT to patients with mild to moderate COVID-19 was associated with clinical improvement with decreasing inflammation and good functional reconstitution.

AAT is an acute phase protein and its production in the liver is induced by inflammatory mediators (22). Clinical data showed that in severe COVID-19 the AAT serum concentration relative to the amount of systemic inflammation is decreased. These data confirm similar findings from the McElvaney group (21) and suggest that reduced activity of AAT could represent a host factor in the modulation of the course of COVID-19 due to the anti-inflammatory properties of AAT (12, 13).

In addition to its anti-inflammatory activities, AAT inhibits SARS-CoV-2 replication in a cell-based human model of COVID-19 in airway organoids, although with modest efficiency. Antiviral activity is likely due to blockade of TMPRSS2 as shown by the decreased broad serine-protease activity of cell supernatants. TMPRSS2 has been shown earlier to activate SARS-CoV-2 and several other viral respiratory pathogens (23). Several other studies have suggested a potential role of AAT in COVID-19. Two studies showed AAT-dependent inhibition of entry of pseudotyped VSV or SARS-CoV-2 into airway cells (24-26). Another study could not detect a significant effect of AAT in cell-based SARS-CoV-2 infection models (27). Differences within the cell models likely contribute to this variation.

We treated 9 COVID-19 patients with AAT, and all patients showed clinical improvement within several days associated with full functional recovery, decreasing viral load and reduced inflammation. In matched controls, four patients died and the decrease of serum CRP was delayed as compared to the treatment group. Based on the small number of patients it is difficult to postulate a significant treatment effect. Other studies showed that between 21.1 and 24.5 % of the patients hospitalized deteriorated and died (28-30).

AAT might represent a good candidate for rapid development for clinical use in COVID-19. The safety data for intravenous and inhaled treatment for AATD—based COPD and other conditions are favorable (20). AAT application might be beneficial in two settings in COVID-19: The one is early disease stages, in which inhaled therapy could decrease local viral load and local inflammation. This type of therapy could also be applied in an outpatient setting. The other setting might be application in severe COVID-19, in which a systemic inflammatory response could be modified by combined systemic and inhaled AAT application.

This study has limitations that need to be addressed, first of all the lack of data from a controlled trial and the missing deep mechanistic characterization of the mode of action of AAT. These items are beyond this report, which shows that AAT has anti-SARS-CoV-2 activity, is well tolerated in patients with COVID-19 and together with its anti-inflammatory properties can be considered a promising candidate for treatment of COVID-19 that should be further evaluated in a randomized controlled trial for inhaled and / or intravenous treatment.

## Data Availability

The data collected for the study, including individual participant data and a data dictionary defining each field in the set, will be made available to others upon reasonable request. The data that will be made available include anonymized clinical data and data dictionaries. Other related documents are available, including the study protocol, statistical analysis plan, informed consent. The data will be available with publication through the email of the corresponding author. Data will be shared after approval by the author group.

## Acknowledgments

The authors thank Nory Astrid Guzman and Susanne Geyer for excellent work as study nurses and Anja Honecker, Andreas Kamyschnikow, and Victoria Weinhold for excellent laboratory work. This work was supported by grants of the Rolf M. Schwiete Stiftung, the Saarland University, BMBF and The States of Saarland and Lower Saxony.

---

**Table S1:** Characteristics of the patients treated with AAT.

| Patient number | Sex | Age | Respiratory status at inclusion | Treatment |
| --- | --- | --- | --- | --- |
| AAT treatment |  |  |  |  |
| 02-07, 02-06 | male | 46 - 54 | 2, 3 | i.v. + inhal. |
| 02-08, 02-09, 02-05 | female | 54 - 79 | 1, 2, 2 | i.v. + inhal. |
| 02-02, '02-03, '02-04 | male | 58 - 72 | 1, 4, 1 | inhal. |
| 02-01 | female | > 50 | 2 | inhal. |
| Means (+/- SD) | - | 60.89<br>(11.06) | 2.00 (1.00) | - |
| Control patients |  |  |  |  |
| Means (+/- SD) | 12 male /<br>6 female | 61.17<br>(10.08) | 2.11 (1.64) | - |

Respiratory status: 1 = spontaneous, 2 = oxygen, 3 = NIV, 4 = high flow oxygen, 5 = invasive, 6 = ECMO; i.v. = intravenous treatment, inhal. = inhaled treatment.

## References

1. Coronaviridae Study Group of the International Committee on Taxonomy of V. The species Severe acute respiratory syndrome-related coronavirus: classifying 2019-nCoV and naming it SARS-CoV-2. Nat Microbiol 2020; 5: 536–544.

2. Beigel JH, Tomashek KM, Dodd LE, Mehta AK, Zingman BS, Kalil AC, Hohmann E, Chu HY, Luetkemeyer A, Kline S, Lopez de Castilla D, Finberg RW, Dierberg K, Tapson V, Hsieh L, Patterson TF, Paredes R, Sweeney DA, Short WR, Touloumi G, Lye DC, Ohmagari N, Oh MD, Ruiz-Palacios GM, Benfield T, Fatkenheuer G, Kortepeter MG, Atmar RL, Creech CB, Lundgren J, Babiker AG, Pett S, Neaton JD, Burgess TH, Bonnett T, Green M, Makowski M, Osinusi A, Nayak S, Lane HC, Members A-SG. Remdesivir for the Treatment of Covid-19 -Final Report. N Engl J Med 2020; 383: 1813–1826.

3. Group RC, Horby P, Lim WS, Emberson JR, Mafham M, Bell JL, Linsell L, Staplin N, Brightling C, Ustianowski A, Elmahi E, Prudon B, Green C, Felton T, Chadwick D, Rege K, Fegan C, Chappell LC, Faust SN, Jaki T, Jeffery K, Montgomery A, Rowan K, Juszczak E, Baillie JK, Haynes R, Landray MJ. Dexamethasone in Hospitalized Patients with Covid-19 -Preliminary Report. N Engl J Med 2020.

4. Salama C, Han J, Yau L, Reiss WG, Kramer B, Neidhart JD, Criner GJ, Kaplan-Lewis E, Baden R, Pandit L, Cameron ML, Garcia-Diaz J, Chávez V, Mekebeb-Reuter M, Lima de Menezes F, Shah R, González-Lara MF, Assman B, Freedman J, Mohan SV. Tocilizumab in Patients Hospitalized with Covid-19 Pneumonia. N Engl J Med 2020.

5. Kim PS, Read SW, Fauci AS. Therapy for Early COVID-19: A Critical Need. JAMA 2020; 324: 2149–2150.

6. Hoffmann M, Kleine-Weber H, Schroeder S, Kruger N, Herrler T, Erichsen S, Schiergens TS, Herrler G, Wu NH, Nitsche A, Muller MA, Drosten C, Pohlmann S. SARS-CoV-2 Cell Entry Depends on ACE2 and TMPRSS2 and Is Blocked by a Clinically Proven Protease Inhibitor. Cell 2020.

7. Heurich A, Hofmann-Winkler H, Gierer S, Liepold T, Jahn O, Pohlmann S. TMPRSS2 and ADAM17 cleave ACE2 differentially and only proteolysis by TMPRSS2 augments entry driven by the severe acute respiratory syndrome coronavirus spike protein. J Virol 2014; 88: 1293–1307.

8. Daly JL, Simonetti B, Klein K, Chen KE, Williamson MK, Anton-Plagaro C, Shoemark DK, Simon-Gracia L, Bauer M, Hollandi R, Greber UF, Horvath P, Sessions RB, Helenius A, Hiscox JA, Teesalu T, Matthews DA, Davidson AD, Collins BM, Cullen PJ, Yamauchi Y. Neuropilin-1 is a host factor for SARS-CoV-2 infection. Science 2020; 370: 861–865.

9. Berlin DA, Gulick RM, Martinez FJ. Severe Covid-19. N Engl J Med 2020.

10. Schulte-Schrepping J, Reusch N, Paclik D, Bassler K, Schlickeiser S, Zhang B, Kramer B, Krammer T, Brumhard S, Bonaguro L, De Domenico E, Wendisch D, Grasshoff M, Kapellos TS, Beckstette M, Pecht T, Saglam A, Dietrich O, Mei HE, Schulz AR, Conrad C, Kunkel D, Vafadarnejad E, Xu CJ, Horne A, Herbert M, Drews A, Thibeault C, Pfeiffer M, Hippenstiel S, Hocke A, Muller-Redetzky H, Heim KM, Machleidt F, Uhrig A, Bosquillon de Jarcy L, Jurgens L, Stegemann M, Glosenkamp CR, Volk HD, Goffinet C, Landthaler M, Wyler E, Georg P, Schneider M, Dang-Heine C, Neuwinger N, Kappert K, Tauber R, Corman V, Raabe J, Kaiser KM, Vinh MT, Rieke G, Meisel C, Ulas T, Becker M, Geffers R, Witzenrath M, Drosten C, Suttorp N, von Kalle C, Kurth F, Handler K, Schultze JL, Aschenbrenner AC, Li Y, Nattermann J, Sawitzki B, Saliba AE, Sander LE, Deutsche C-OI. Severe COVID-19 Is Marked by a Dysregulated Myeloid Cell Compartment. Cell 2020; 182: 1419–1440 e1423.

11. Strnad P, McElvaney NG, Lomas DA. Alpha1-Antitrypsin Deficiency. N Engl J Med 2020; 382: 1443–1455.

12. Schuster R, Motola-Kalay N, Baranovski BM, Bar L, Tov N, Stein M, Lewis EC, Ayalon M, Sagiv Y. Distinct anti-inflammatory properties of alpha1-antitrypsin and corticosteroids reveal unique underlying mechanisms of action. Cell Immunol 2020; 356: 104177.

13. McElvaney OJ, O’Connor E, McEvoy NL, Fraughan DD, Clarke J, McElvaney OF, Gunaratnam C, O’Rourke J, Curley GF, McElvaney NG. Alpha-1 antitrypsin for cystic fibrosis complicated by severe cytokinemic COVID-19. Journal of cystic fibrosis : official journal of the European Cystic Fibrosis Society 2020.

14. Stolk J, Tov N, Chapman KR, Fernandez P, MacNee W, Hopkinson NS, Piitulainen E, Seersholm N, Vogelmeier CF, Bals R, McElvaney G, Stockley RA. Efficacy and safety of inhaled alpha1-antitrypsin in patients with severe alpha1-antitrypsin deficiency and frequent exacerbations of COPD. Eur Respir J 2019; 54.

15. Chapman KR, Stockley RA, Dawkins C, Wilkes MM, Navickis RJ. Augmentation therapy for alpha1 antitrypsin deficiency: a meta-analysis. Copd 2009; 6: 177–184.

16. Esumi M, Ishibashi M, Yamaguchi H, Nakajima S, Tai Y, Kikuta S, Sugitani M, Takayama T, Tahara M, Takeda M, Wakita T. Transmembrane serine protease TMPRSS2 activates hepatitis C virus infection. Hepatology 2015; 61: 437–446.

17. Sprott RF, Ritzmann F, Langer F, Yao Y, Herr C, Kohl Y, Tschernig T, Bals R, Beisswenger C. Flagellin shifts 3D bronchospheres towards mucus hyperproduction. Respiratory research 2020; 21: 222.

18. Kleine-Weber H, Elzayat MT, Wang L, Graham BS, Muller MA, Drosten C, Pohlmann S, Hoffmann M. Mutations in the Spike Protein of Middle East Respiratory Syndrome Coronavirus Transmitted in Korea Increase Resistance to Antibody-Mediated Neutralization. J Virol 2019; 93.

19. Berger Rentsch M, Zimmer G. A vesicular stomatitis virus replicon-based bioassay for the rapid and sensitive determination of multi-species type I interferon. PloS one 2011; 6: e25858.

20. Stolk J, Tov N, Chapman KR, Fernandez P, MacNee W, Hopkinson NS, Piitulainen E, Seersholm N, Vogelmeier CF, Bals R, McElvaney G, Stockley RA. Efficacy and safety of inhaled alpha-1-antitrypsin in patients with severe alpha-1-antitrypsin deficiency and frequent exacerbations of Chronic Obstructive Pulmonary Disease. Eur Respir J 2019.

21. McElvaney OJ, McEvoy NL, McElvaney OF, Carroll TP, Murphy MP, Dunlea DM, Ni Choileain O, Clarke J, O’Connor E, Hogan G, Ryan D, Sulaiman I, Gunaratnam C, Branagan P, O’Brien ME, Morgan RK, Costello RW, Hurley K, Walsh S, de Barra E, McNally C, McConkey S, Boland F, Galvin S, Kiernan F, O’Rourke J, Dwyer R, Power M, Geoghegan P, Larkin C, O’Leary RA, Freeman J, Gaffney A, Marsh B, Curley GF, McElvaney NG. Characterization of the Inflammatory Response to Severe COVID-19 Illness. Am J Respir Crit Care Med 2020; 202: 812–821.

22. Perlmutter DH, May LT, Sehgal PB. Interferon beta 2/interleukin 6 modulates synthesis of alpha 1-antitrypsin in human mononuclear phagocytes and in human hepatoma cells. J Clin Invest 1989; 84: 138–144.

23. Hatesuer B, Bertram S, Mehnert N, Bahgat MM, Nelson PS, Pohlmann S, Schughart K. Tmprss2 is essential for influenza H1N1 virus pathogenesis in mice. PLoS Pathog 2013; 9: e1003774.

24. Oguntuyo KY, Stevens CS, Siddiquey MN, Schilke RM, Woolard MD, Zhang H, Acklin JA, Ikegame S, Hung CT, Lim JK, Cross RW, Geisbert TW, Ivanov SS, Kamil JP, Lee B. In plain sight: the role of alpha-1-antitrypsin in COVID-19 pathogenesis and therapeutics. bioRxiv 2020.

25. Azouz NP, Klingler AM, Callahan V, Akhrymuk IV, Elez K, Raich L, Henry BM, Benoit JL, Benoit SW, Noe F, Kehn-Hall K, Rothenberg ME. Alpha 1 Antitrypsin is an Inhibitor of the SARS-CoV-2-Priming Protease TMPRSS2. bioRxiv 2020.

26. Wettstein L, Conzelmann C, Müller JA, Weil T, Groß R, Hirschenberger M, Seidel A, Klute S, Zech F, Bozzo CP, Preising N, Fois G, Lochbaum R, Knaff P, Mailänder V, Ständker L, Thal DR, Schumann C, Stenger S, Kleger A, Lochnit G, Sparrer K, Kirchhoff F, Frick M, Münch J. Alpha-1 antitrypsin inhibits SARS-CoV-2 infection. bioRxiv 2020: 2020.2007.2002.183764.

27. Bojkova D, Bechtel M, McLaughlin KM, McGreig JE, Klann K, Bellinghausen C, Rohde G, Jonigk D, Braubach P, Ciesek S, Munch C, Wass MN, Michaelis M, Cinatl J, Jr. Aprotinin Inhibits SARS-CoV-2 Replication. Cells 2020; 9.

28. Richardson S, Hirsch JS, Narasimhan M, Crawford JM, McGinn T, Davidson KW, the Northwell C-RC, Barnaby DP, Becker LB, Chelico JD, Cohen SL, Cookingham J, Coppa K, Diefenbach MA, Dominello AJ, Duer-Hefele J, Falzon L, Gitlin J, Hajizadeh N, Harvin TG, Hirschwerk DA, Kim EJ, Kozel ZM, Marrast LM, Mogavero JN, Osorio GA, Qiu M, Zanos TP. Presenting Characteristics, Comorbidities, and Outcomes Among 5700 Patients Hospitalized With COVID-19 in the New York City Area. JAMA 2020; 323: 2052–2059.

29. Argenziano MG, Bruce SL, Slater CL, Tiao JR, Baldwin MR, Barr RG, Chang BP, Chau KH, Choi JJ, Gavin N, Goyal P, Mills AM, Patel AA, Romney MS, Safford MM, Schluger NW, Sengupta S, Sobieszczyk ME, Zucker JE, Asadourian PA, Bell FM, Boyd R, Cohen MF, Colquhoun MI, Colville LA, de Jonge JH, Dershowitz LB, Dey SA, Eiseman KA, Girvin ZP, Goni DT, Harb AA, Herzik N, Householder S, Karaaslan LE, Lee H, Lieberman E, Ling A, Lu R, Shou AY, Sisti AC, Snow ZE, Sperring CP, Xiong Y, Zhou HW, Natarajan K, Hripcsak G, Chen R. Characterization and clinical course of 1000 patients with coronavirus disease 2019 in New York: retrospective case series. BMJ 2020; 369: m1996.

30. Karagiannidis C, Mostert C, Hentschker C, Voshaar T, Malzahn J, Schillinger G, Klauber J, Janssens U, Marx G, Weber-Carstens S, Kluge S, Pfeifer M, Grabenhenrich L, Welte T, Busse R. Case characteristics, resource use, and outcomes of 10 021 patients with COVID-19 admitted to 920 German hospitals: an observational study. The Lancet Respiratory medicine 2020; 8: 853–862.

